# Clinical Features and Vaccine Efficacy Analysis of COVID-19 patients in a Chongqing Shelter hospital in 2022

**DOI:** 10.1101/2023.09.21.23295669

**Authors:** YiNing Luo, Mei Zhao, Xingyu Zhao, YanWen Qiao, YanXia Gao, TingTing Wu, Weiwei Liu, Yi Ren

## Abstract

**Background:** The long-term impact of coronavirus disease 2019 (COVID-19) on many aspects of society emphasizes the necessity of vaccination and nucleic acid conversion time as markers of prevention and diagnosis. However, little research has been conducted on the immunological effects of vaccines and the influencing factors of virus clearance. Epidemiological characteristics and factors related to disease prognosis and nucleic acid conversion time need to be explored.

**Design and participants:** We reviewed published documentation to create an initial draft. The data were then statistically evaluated to determine their link. Given that a Chongqing shelter hospital is typical in terms of COVID-19 patients receiving hospital management and treatment effects, a retrospective analysis was conducted on 4,557 cases of COVID-19 infection in a shelter hospital in Chongqing in December 2022, which comprised 2,291 males and 2,266 females. The variables included age, medical history, nucleic acid conversion time, vaccination status, and clinical symptoms.

**Results:** Univariate survival analysis using the Log-rank test (P < 0.05) showed that factors such as age significantly affected nucleic acid conversion time. COX regression analysis indicated a significant association between a history of hypertension and nucleic acid conversion time, which had a hazard ratio of 0.897 (95% CI: 0.811–0.992). A statistically significant difference was observed between vaccinated and unvaccinated infected individuals in terms of the presence of symptoms such as cough and sensory system manifestations (P < 0.05).

**Conclusion:** The effect of vaccination against COVID-19 on symptoms such as coughing, nasal congestion, muscle aches, runny nose, and sensory system symptoms in COVID-19 patients was determined. Typical symptoms, such as runny nose, were generally higher in vaccinated than in unvaccinated ones; previous hypertension was an influential factor in nucleic acid conversion time in patients with COVID-19 infection.

**STRENGTHS AND LIMITATIONS:** This study clarifies the advantages of vaccination by comparing the symptom development between patients who had received vaccinations and those who had not.

This research suggests potential future study directions, such as investigating the impact of pre-existing diseases like hypertension on viral clearance time and assessing vaccine efficacy and safety in certain demographic groups.

This work investigated data from 4,557 coronavirus disease 2019 patients admitted to a shelter hospital, which offered significant insights into patients’ clinical features and outcomes over a defined time span.

This study has several flaws because a retrospective analysis method was used and not all confounding variables that might have affected the results were appropriately controlled.

The overall research sample may not be representative of other communities because it was drawn from a shelter hospital in Chongqing, and the types of immunizations used were not disclosed. These factors might have had an impact on the precision and extent of the research findings.

## Introduction

Coronavirus disease 2019 (COVID-19), which is caused by the novel coronavirus, is a highly contagious infectious disease that is primarily transmitted through respiratory droplets and affects individuals of all age groups and diverse social backgrounds^1^. Common clinical manifestations include dry throat, sore throat, cough, fever, and fatigue, with some patients experiencing additional symptoms such as loss or reduction in taste and smell, nasal congestion, diarrhea, vomiting, and chest radiographic changes^2^. The World Health Organization (WHO) assessment reported that, during the convalescence of COVID-19 infection, approximately 10%–20% of patients may experience persistent symptoms for several months, which are characterized by prolonged or recurrent manifestations^3^. Furthermore, the WHO data released on April 27, 2023 indicated that the global reported cases of COVID-19 were more than 760 million, with over 6.9 million deaths^4^. This pandemic has profoundly impacted the quality of life, public health systems, and society governance worldwide, which requires urgent development of appropriate strategies to combat the virus.

Therapeutic effect of COVID-19 is affected by various factors. At present, the negative conversion time of nucleic acid and shelter hospital are the important affecting factors of the therapeutic effect. The period of nucleic acid negative conversion is an essential criterion for the discharge of COVID-19 patients from isolation. However, research on this problem in China and worldwide is still restricted and lacks consensus due to practical constraints. Current research has predominantly focused on underlying medical conditions^5–7^ and immunological aspects^8–10^. In addition, vaccination stands as a major measure for preventing COVID-19 infection and has been extensively implemented across the globe^11,12^, and it significantly reduces hospitalizations, severe cases, and deaths caused by the disease^13^. Existing research has mostly focused on the influencing factors of vaccination, such as national population size^14^, ethnicity, educational level^15^, and perception of disease threat^16^. However, research on the immune protective effects of vaccination remains limited.

Shelter hospitals, as a critical measure to combat virus transmission, are a unique medical response system with Chinese characteristics, and they were developed by the Chinese government in response to the outbreak of COVID-19 in Hubei Province, particularly in Wuhan City^17^. The construction of shelter hospitals, which was initially implemented as an emergency initiative in response to the COVID-19 transmission in Wuhan, has been conducted in three batches. This initiative played a vital role in alleviating the situation in Wuhan and ultimately achieving success in the battle against the pandemic^18^. On account of its vast population of patients and being at the front line of treating COVID-19 patients, Wuhan has relatively completed first-hand clinical data of COVID-19 patients. On average, one out of every five patients in Wuhan in 2020 was treated in a shelter hospital. This facility has played an irreplaceable key role in promoting the treatment of pneumonia caused by COVID-19^19^.

In view of the outstanding performance of shelter hospitals in the prevention and control of the Wuhan epidemic, other regions in China, such as Chongqing, have also set up shelter hospitals based on their own epidemic prevention and control conditions. November to the end of December 2022 is a critical period for the prevention and control of COVID-19 in Chongqing. Given that the situation of epidemic prevention and control in Chongqing is the most severe, taking reasonable and effective prevention and control measures is important. Chongqing has set up shelter hospitals to meet the needs of COVID-19 prevention and control. During this period, more than 4,000 COVID-19 patients were admitted to the hospital in time. Effective clinical treatment has fully relieved the medical pressure in Chongqing and further curbed the spread of COVID-19, which makes important contributions to the successful prevention and control of the epidemic.

Shelter hospitals, as an important defense line to effectively curb the spread of the novel coronavirus, have increasingly become the research focus of scholars in related fields. In recent years, researchers have found that most of the large-scale shelter hospitals have short preparation time, great difficulties in admission and treatment, and high potential cost of integration. Given this phenomenon, academics are now focusing on optimizing the management efficiency of shelter hospitals^20,21^, the hardware equipment^22,23^, and the evaluation of clinical treatment effect and its influencing factors^24^. Among them, vaccination and nucleic acid clearance time are important affecting factors of the clinical treatment effect of COVID-19 patients in shelter hospitals, which need to be studied to better play the role of shelter hospitals. However, studies on the influencing factors of nucleic acid conversion time, the immunological protection of vaccines, and the impact of vaccination on patients in shelter hospitals remain relatively scarce domestically and internationally.

This study aims to analyze the impact of vaccination on the clinical characteristics of COVID-19 patients in a shelter hospital in Chongqing from November to December 2022. It also aims to explore the immune protective effect of vaccine and the influencing factors of the negative conversion time of nucleic acid by collecting the clinical data of COVID-19 patients. Moreover, it aims to provide a more reliable basis for the application of vaccine in the prevention and control of virus transmission and a more accurate reference for clinical treatment decisions.

## Methods

### Setting

The research was conducted from November 2022 to January 2023. It was performed in collaboration with a shelter hospital in Chongqing, which is a medical school in Chongqing.

### Data Collection

A retrospective analysis was conducted on 4,557 confirmed COVID-19 patients who were admitted to a certain shelter hospital in Chongqing from November to the end of December 2022. The investigators collected patients’ general information, pre-infection vaccination status (self-reported), post-infection clinical symptoms, and past medical history (diagnosed by a doctor), with a sample validity rate of 100%. The inclusion criteria were as follows: (1) all patients met the diagnostic criteria for COVID-19 according to the *Scheme for Diagnosis and Treatment of 2019 Novel Coronavirus Pneumonia (The 10th Trial Edition)*^25^ issued by the National Health Commission of China; (2) complete clinical data were available. The exclusion criteria were as follows: (1) deficient nucleic acid conversion time and vaccination status; (2) patients with no follow-up data due to transfer or death. The study parameters included participants’ age, gender, history of hypertension, coronary heart disease, fever, cough, vaccination status, and nucleic acid conversion time.

### Clinical Signs of Infection

On the basis of the Scheme for Diagnosis and Treatment of 2019 Novel Coronavirus Pneumonia (The 10th Trial Edition) jointly released by the National Health Commission and other relevant departments in China, this study primarily explored clinical signs of infection such as cough, sore throat, fever, muscle pain, nasal congestion, rhinorrhea, sensory system manifestations (reduced or loss of smell and taste), and gastrointestinal symptoms (anorexia, diarrhea, nausea, and bloating) in COVID-19 patients.

### Statistical Methods

Categorical variables were presented as percentages or frequencies, and average nucleic acid conversion time was expressed as mean ± standard deviation. First, survival analysis models were used to explore the influencing factors of the time of nucleic acid conversion in COVID-19 patients, such as age, history of hypertension and diabetes, occurrence of typical COVID-19 symptoms after infection, and vaccination status. Single-factor analysis was conducted using the Log-rank test, and variables with P < 0.05 were included in the Cox proportional hazard model. By estimating hazard ratios (HRs) and 95% confidence intervals (CIs), we determined the affecting factors of the time a patient’s nucleic acid test takes to turn negative. Simultaneously, the Chi-square test was used to investigate the immunological protection of vaccination on typical symptoms of COVID-19 among infected individuals. Given that medical history might influence the presence of symptoms in COVID-19 patients, 3,822 cases without any medical history were selected from the total of 4,557 infected individuals for this study. Data cleaning and statistical analysis were performed using SPSS 26.0 software, and statistical significance was set at bilateral P < 0.05 for all analyses.

## Results

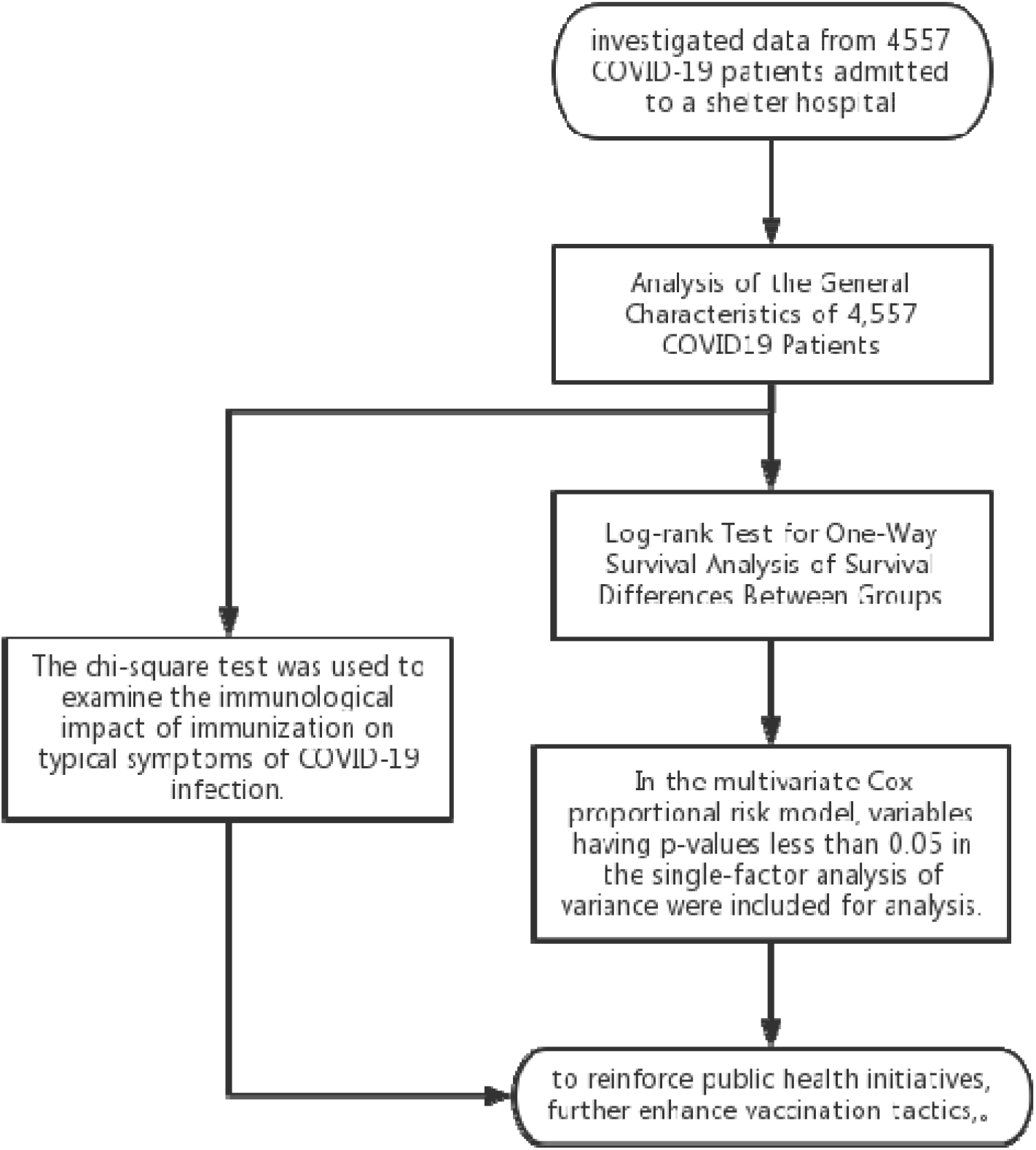

### Demographic Characteristics of COVID-19 Patients

A total of 4,557 confirmed COVID-19 patients were included based on the diagnostic criteria of the *Scheme for Diagnosis and Treatment of 2019 Novel Coronavirus Pneumonia (The 10th Trial Edition)*^25^. Among them, asymptomatic infections accounted for 22.5% of the cases. The age group mainly ranged from 18 years to 65 years, which comprised approximately 84.3% of the cases. About 16.1% of the infected individuals had a history of underlying medical conditions. The average nucleic acid conversion time was 6.94 (±2.60) days, and approximately 94.3% of the infected patients had received vaccination from any vaccine manufacturer. Specific details are shown in Table 1.

**Table 1.** General Characteristics Analysis of 4,557 COVID19 Patients.

|  | Characteristics | Number of Patients (n=4,557) |
| --- | --- | --- |
| **Gender (cases)** | Male | 2291 (50.3%) |
|  | Female | 2266 (49.7%) |
| **Age Group (cases)** | 0–17 years | 357 (7.8%) |
|  | 18–65 years | 3843 (84.3%) |
|  | 66–79 years | 345 (7.6%) |
|  | ≥80 years | 12 (0.3%) |
| **History of Underlying Medical Conditions** | Yes | 735 (16.1%) |
|  | No | 3822 (83.8%) |
| **Average Nucleic Acid Conversion Time (days)** | / | 6.94 (±2.60) |
| **Symptomatic Cases** | Yes | 3530 (77.5%) |
|  | No | 1027 (22.5%) |
| **Vaccination Status** | Vaccinated | 4299 (94.3%) |
|  | Unvaccinated | 258 (5.7%) |

### Factors Associated with Nucleic Acid Conversion Time

#### Single-Factor Survival Analysis using Log-rank Test

The Log-rank test results showed that age, history of hypertension, history of surgery, and history of allergies exhibited statistically significant differences among different groups (P < 0.05), as presented in Table 2.

**Table 2.**
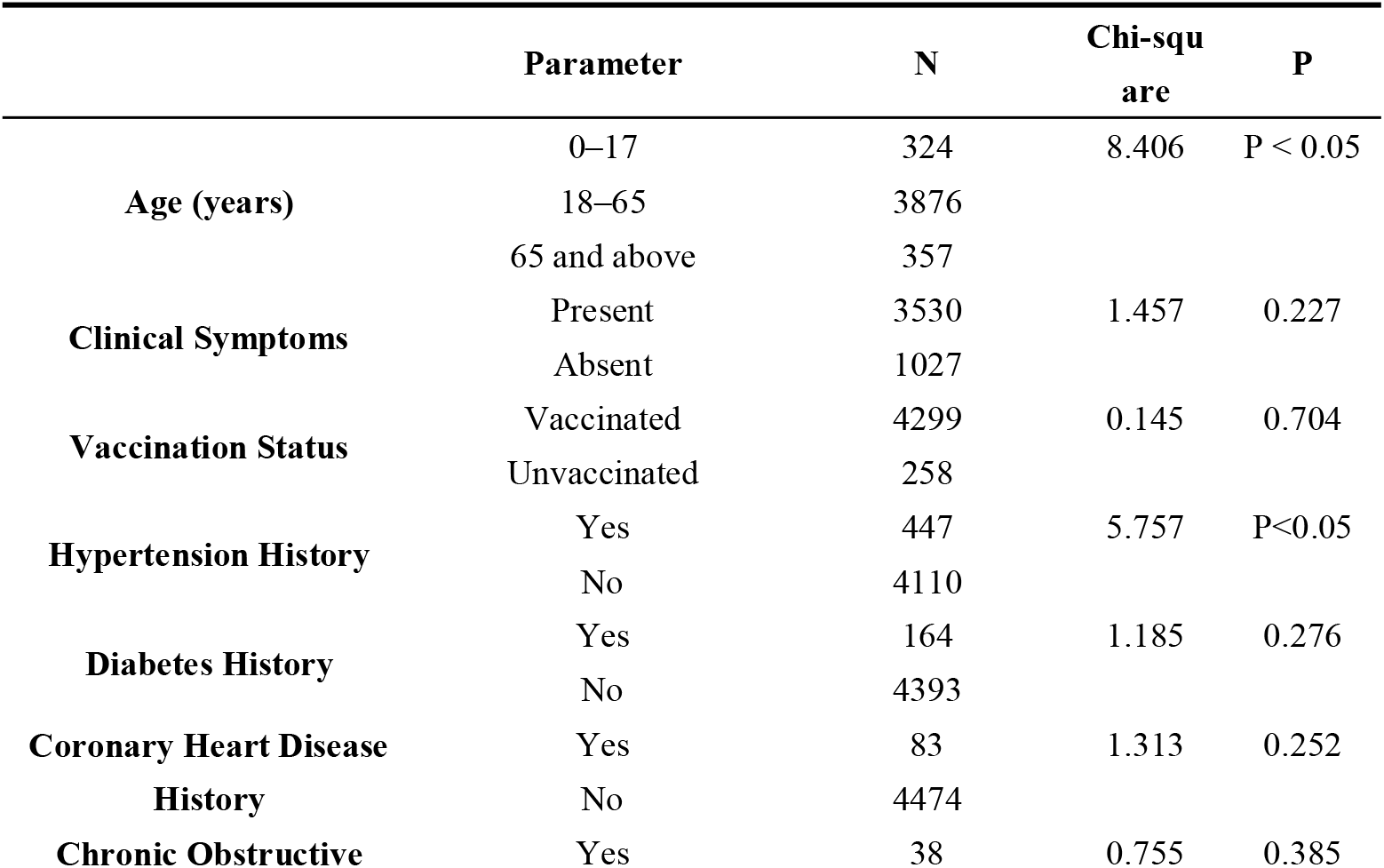
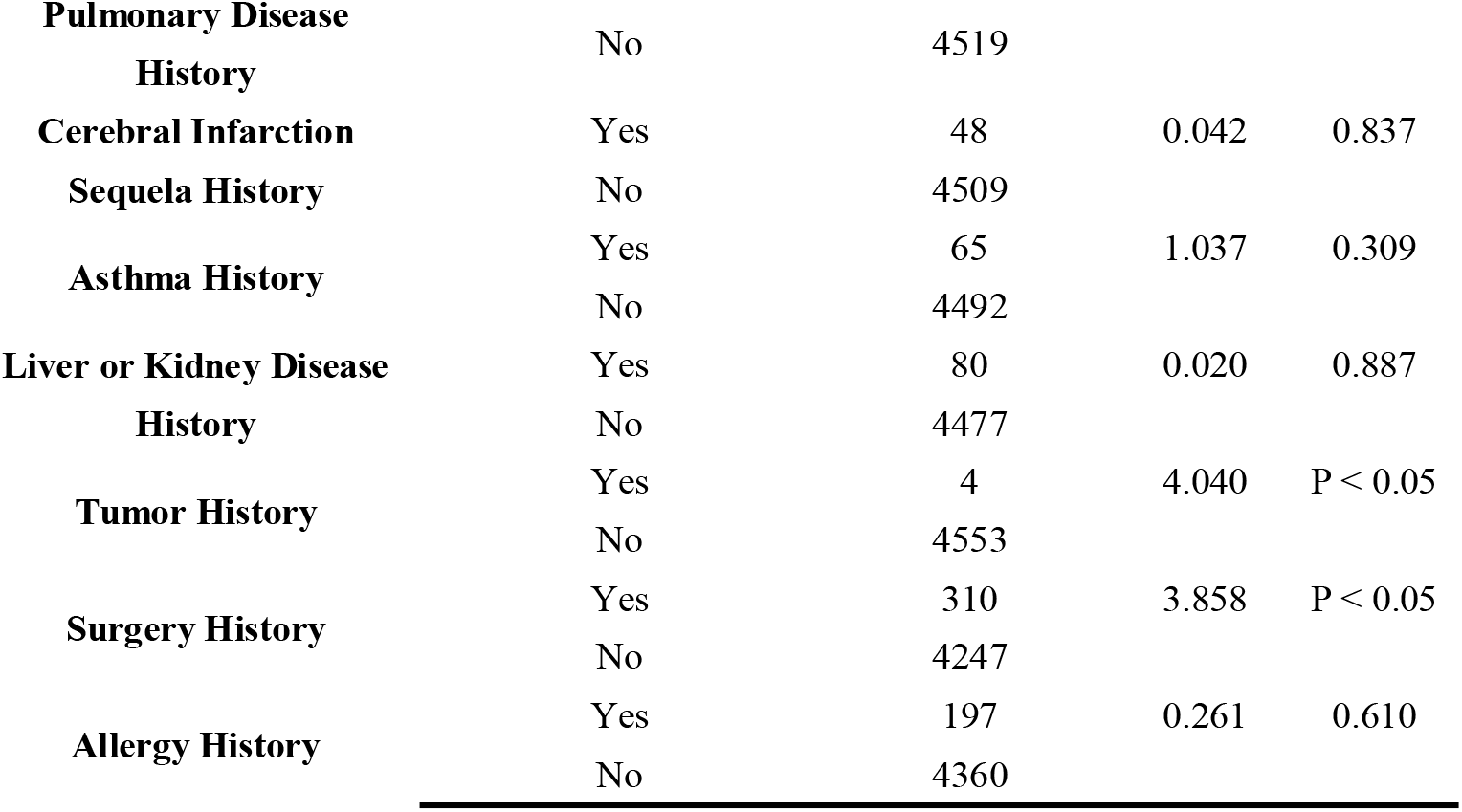
Results of Log-rank Test Analysis (n=4,557)

### Multivariable Cox Proportional Hazard Model

The variables with a P value less than 0.05 in the single-factor analysis were included in the multivariable Cox proportional hazard model. The results showed that a history of hypertension in infected individuals was negatively correlated with nucleic acid conversion time, with an HR of 0.897 (95% CI: 0.811–0.992). This finding suggests that individuals with a history of hypertension had a relatively shorter time to achieve viral clearance, as presented in Table 3.

**Table 3.** Multivariable Cox Regression Analysis (n=4,557)

|  | **b** | **SE** | **Wald x<sup>2</sup>** | **P** | **HR** | **95%CI** |
| --- | --- | --- | --- | --- | --- | --- |
| **Hypertension** | −0.108 | 0.051 | 4.461 | P < 0.05 | 0.897 | (0.811, 0.992) |
| **Age** | 0.046 | 0.038 | 1.427 | 0.232 | 1.047 | (0.971, 1.128) |
| **Tumor History** | −0.754 | 0.501 | 2.263 | 0.132 | 0.471 | (0.176, 1.256) |
| **Surgery History** | −0.100 | 0.059 | 2.868 | 0.090 | 0.905 | (0.806, 1.016) |

### Analysis of the Immunological Protection of Vaccination on Typical Symptoms of COVID-19 Infection

The Chi-square test findings revealed significant differences (P < 0.05) in symptoms, including cough, nasal congestion, muscle soreness, runny nose, and sensory system symptoms, between vaccinated and unvaccinated individuals. However, no significant differences (P > 0.05) were observed in symptoms on sore throat, fever, and gastrointestinal symptoms. The detailed results are presented in Table 4.

**Table 4.**
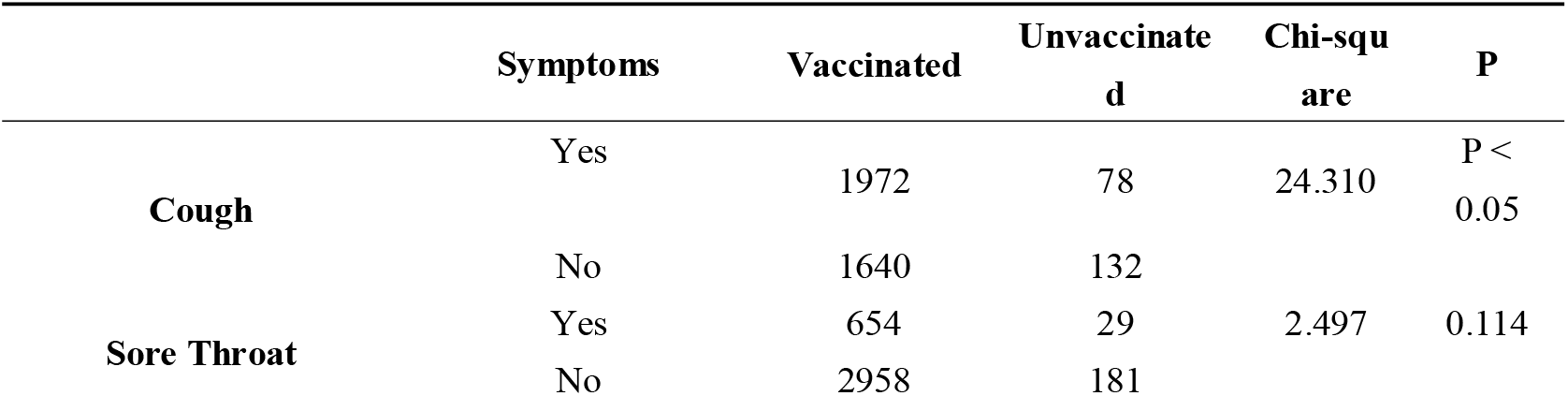
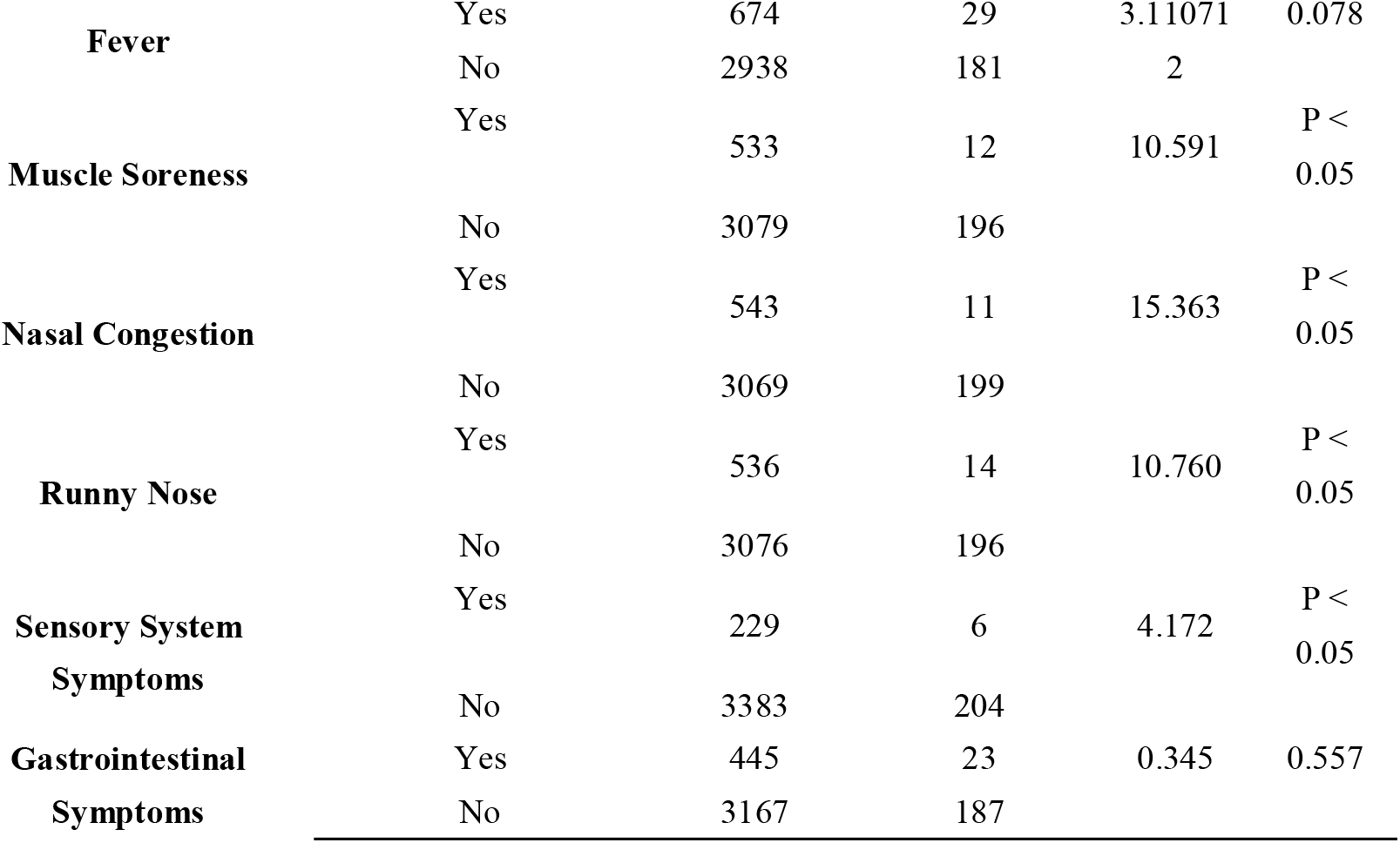
Chi-square Test Results for Vaccination and Typical Symptoms (n=3,822)

## Discussion

On the basis of data from the studied shelter hospital, researchers examined the association between vaccinations and cough, nasal congestion, runny nose, and other common symptoms in COVID-19 patients. The results of this study indicate a significant difference in the development of typical symptoms between vaccinated and unvaccinated individuals. COVID-19 patients who received vaccinations were more likely to exhibit typical symptoms. However, the results of the study by Zhao et al. showed that unvaccinated patients with novel coronavirus infection had a higher probability of experiencing clinical symptoms such as fever, fatigue, and muscle aches and pains than vaccinated patients with novel coronavirus infection^26^.

Numerous studies have shown that vaccination triggers the body’s immune response, which generates specific antibodies and cellular immunity that can reduce the risk of COVID-19 infection, particularly severe cases and fatalities^27,28^. Vaccination also helps decrease the duration of infection and reduces the risk of viral transmission^29,30^. These antibodies can neutralize the virus, prevent further infection of human cells, and assist other immune cells in eliminating the infected virus. The immune response might result in some short-term discomfort, such as fever, fatigue, and muscle soreness, which are normal manifestations of a functioning immune system. However, these reactions are usually mild and resolve quickly. By contrast, individuals who remain unvaccinated may face more severe outcomes once infected with the COVID-19 due to the lack of immune defenses specific to the virus. The virus may have a higher chance to replicate extensively in the body and trigger an inflammatory response. This condition can lead to severe symptoms, including but not limited to palpitations, difficulty breathing, lung damage, and organ failure^25^. Vaccination is thus widely regarded as a safe and effective strategy to protect persons and lessen the effects of COVID-19 infection, which is despite the fact that patients immunized based on the findings of this study were more likely to have moderate symptoms. Some relative research has found that people who have common symptoms after receiving the vaccine actually experience more severe clinical symptoms. It is worth noting that vaccination is aimed at reducing the likelihood of patients suffering from serious illnesses. The immune response stimulated by common symptoms may prevent patients from developing serious diseases.

Although vaccination significantly reduces the risk of infection, the effect is not absolute. Individuals may still contract the virus after vaccination, which could be due to the imperfect efficacy of the vaccine or variations in the immune responses of individuals. Studies have also suggested that new virus variants may challenge the protective efficacy of existing vaccines, and previous vaccines may have significantly reduced efficacy against the Omicron variant^30,31^. Thus, even after vaccination, individuals should continue to follow public health guidelines, such as wearing masks, maintaining social distancing, and practicing good hand hygiene, to further reduce the risk of infection and protect themselves and others from the consequences of virus transmission^25^.

The effect of a history of hypertension on the time of nucleic acid conversion in COVID-19 patients can be evaluated and determined using data obtained from patients in shelter hospitals. Patients with hypertension often have reduced immune function, increased inflammatory responses, and vascular damage. Molecular pathway analyses suggest that hypertension may promote the development of COVID-19 by inducing inflammatory pathways, which increases the risk of severe illness and death in infected patients^32,33^. Studies have also reported that patients with pulmonary arterial hypertension (PAH) have relatively higher rates of severe illness and mortality after COVID-19 infection^34^.

However, the results of this study showed that a history of hypertension in infected individuals is negatively correlated with the nucleic acid conversion time. Infected individuals with a history of hypertension tend to have a relatively shorter time to achieve viral clearance. Notably, therapeutic drugs used for PAH treatment might provide some protective effects through various mechanisms on pulmonary endothelial cells^34^. Referring to the latest recommendations from the European Society of Cardiology and European Respiratory Society^35^, combination therapy of endothelin receptor antagonists, phosphodiesterase 5 inhibitors, and/or proteinoids can be beneficial to patients (62%)^36^.

In hypertensive individuals, angiotensin II receptor antagonists may have an effect on patients with new coronavirus infections in addition to hypertension, which contributes to a faster time to initiate nucleic acid conversion. Animal studies (on mice) have revealed that the expression of angiotensin-converting enzyme (ACE)2 was significantly elevated among patients treated with angiotensin-converting enzyme inhibitors (ACEIs) or angiotensin receptor blockers (ARBs)^37,38^. On the basis of these findings, some researchers suggest that the usage of ACEIs/ARBs, which causes greater levels of ACE2, may enhance COVID-19 infection^39^. Additional epidemiologic studies and prospective trials could be conducted in the future to determine whether the use of these medicines in individuals with and without other clinical reasons for ACEIs/ARBs reduces the rate of severe disease or fatality due to COVID-19. However, convincing evidence that ACEIs/ARBs are useful in COVID-19 patients is lacking^39^.

This study only discusses the theoretical impact of hypertension on the recovery of COVID-19 patients. Future research can further investigate the relationship between hypertension and the immune response to COVID-19 infection through a closer integration of clinical and laboratory data, which provides a basis for developing individualized treatment plans for hypertensive patients.

In the studied shelter hospital, no significant relationship was detected between vaccination and nucleic acid conversion time in COVID-19 patients. According to the research of Li et al., patients who received two doses of vaccine had significantly lower viral loads and shorter times to peak viral loads than unvaccinated patients, and the time to viral clearance was also significantly shorter in these patients^40^. However, the results of the current study showed a significance value of 0.077 between vaccination and nucleic acid conversion time in COVID-19 patients, and the results did not reach the level of statistical significance. Therefore, no significant relationship existed between vaccination and nucleic acid conversion time in COVID-19 patients. We believe that this finding may be related to the difference in strains of the disease to a certain extent. Some studies have also suggested that omicron can weaken the protective effect of the vaccine^41^. In addition to the abovementioned reasons, the management of the shelter hospitals and the patient–physician and patient–patient interactions in the shelter hospitals may impact the relationship between vaccination and nucleic acid conversion time in COVID-19 patients. The relationship between the two components still deserves further in-depth exploration. Therefore, this relationship can be explored in greater depth during subsequent studies to better address the challenges posed by epidemic prevention and control.

## Conclusion

This retrospective analysis investigated the clinical characteristics and vaccine efficacy of 4,557 COVID-19 patients admitted to a shelter hospital in Chongqing from November to December 2022. The results emphasized the importance of vaccines in COVID-19 prevention and provided relevant references for patients with pre-existing hypertension. Subsequent research can further explore the impact mechanism of pre-existing conditions such as hypertension on the time to viral clearance, as well as the effectiveness and safety of vaccines in specific populations. In addition, multi-center studies with cross-regional designs will help improve the generalizability and reliability of research conclusions, which will further enhance vaccination strategies and strengthen public health interventions to jointly combat the spread of COVID-19. In conclusion, vaccination is an effective way to protect people. Thus, a vaccine against the covid-19 deserves more comprehensive research.

## Data Availability

All data generated in this study are available to the authors upon reasonable request.

## Data availability statement

Data are available on reasonable request. Deidentified raw data are available on reasonable request to the corresponding author.

## Ethics Statement

Ethical review and approval were not required for the study on human participants in accordance with the local legislation and institutional requirements. The patients/participants provided their written informed consent to participate in this study.

## Acknowledgments

We thank the patient for her participation to the study.

